# Immortal Time Bias Can Reverse the Estimated Association: A Clone-Censor-Weight Analysis in an EHR-Based HPV Vaccination Target Trial Emulation

**DOI:** 10.64898/2026.04.21.26351413

**Authors:** Tuo Lin, Yongqiu Li, Zhuochao Huang, Yiwen Chen, Toni T. Gui, Yi Guo

**Author notes:** Corresponding author: Tuo Lin, & Yi Guo.

## Abstract

Using an electronic health record-based human papillomavirus vaccination example, we show that immortal time bias can reverse the direction of an estimated association. We link clone-censor-weight analysis to the potential outcome framework, assess censoring-weight stability, and demonstrate dependence-aware inference, providing practical guidance for longitudinal target trial emulation with treatment strategies defined during follow-up.

## 1 Introduction

Target trial emulation (TTE) prespecifies eligibility, treatment strategies, time zero, follow-up, outcomes, and censoring rules to structure electronic health record (EHR) analyses. In longitudinal TTE, using treatment observed after time zero to define groups can assign event-free pretreatment time to those treated later, creating immortal time bias that may distort or reverse the estimated association; baseline adjustment cannot correct this use of future information [1, 2].

The clone-censor-weight (CCW) approach addresses this design problem by assigning a clone of each eligible individual to every strategy at time zero. Clones are censored when observed treatment becomes incompatible with the assigned strategy, and uncensored records are weighted for this artificial censoring [3, 4]. This cloning step mirrors the potential outcome framework: each individual is represented under every treatment strategy being compared [5].

Several practical issues remain: TTE elements and grace-period data processing must be pre-specified; artificial censoring distinguished from administrative censoring; inverse probability of censoring weighting (IPCW) evaluated using diagnostics and sensitivity analyses; and dependence between clones accounted for in variance estimation. Stabilized weights may reduce extreme-weight influence but do not replace these checks. The weighted Cox hazard ratio is a model-based per-protocol contrast and should not automatically be interpreted as a causal effect. Using an EHR-based HPV vaccination comparison, we illustrate these issues and show how correcting immortal time bias can reverse the estimated association.

## 2 Methods

We conducted a retrospective TTE using the UF Health Integrated Data Repository from January 1, 2012, through February 28, 2025 (Figure 1). The complete target trial protocol is provided in Multimedia Appendix Table S1. Eligible individuals were female, aged 15-26 years at their first 9vHPV dose, had a health care encounter during the prior year and after the 12-month grace period, and met prespecified dosing-interval criteria. We excluded individuals with baseline HIV, immunosuppressive conditions, hysterectomy, pregnancy, sexually transmitted infection, high-risk HPV positivity, abnormal cervical cytology or histology, or cervical cancer.

**Figure 1:**
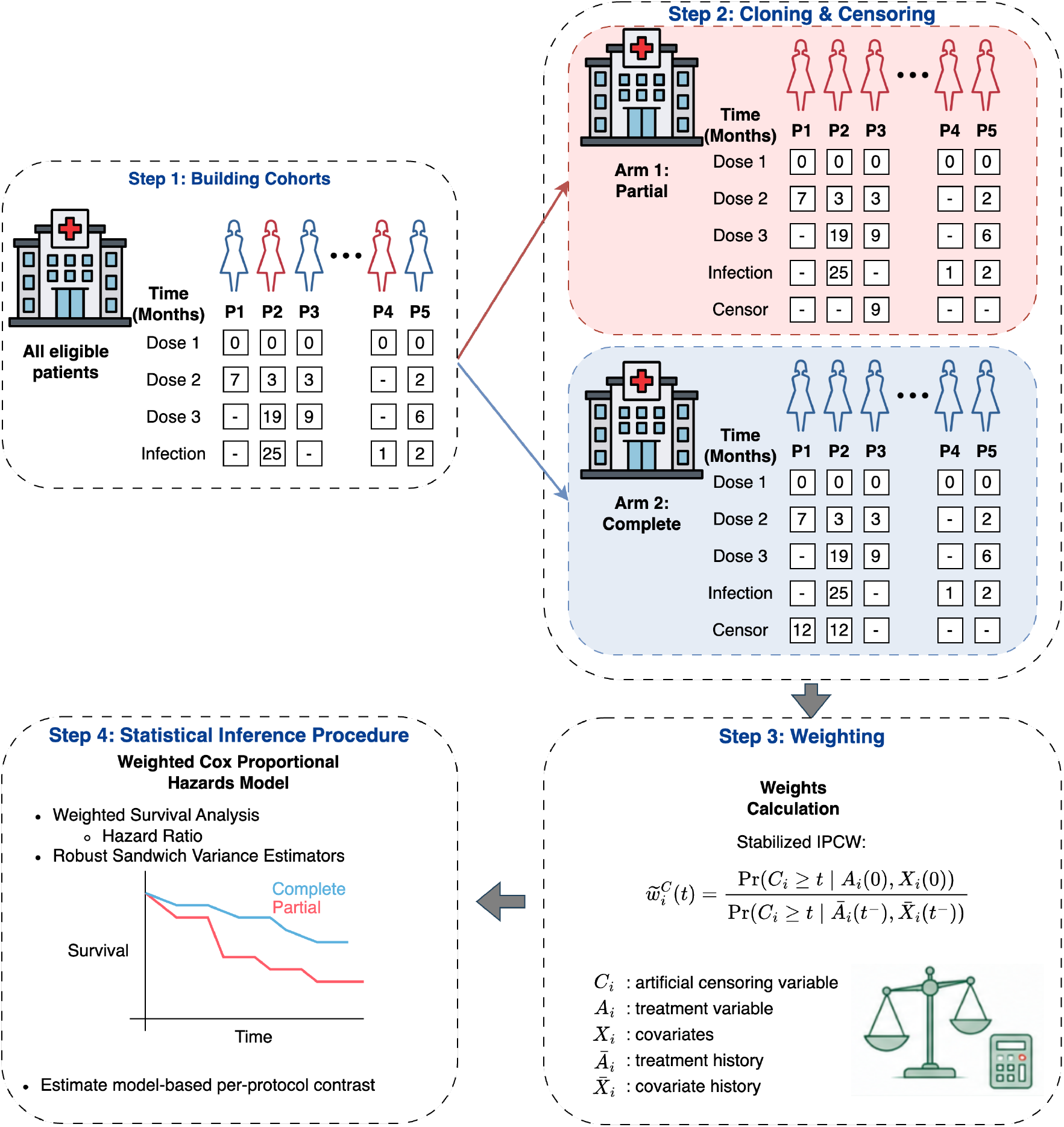
Overview of the clone-censor-weight framework in a retrospective TTE comparing HPV vaccination strategies.

Time zero was the first 9vHPV dose. Each individual was cloned and assigned to both strategies: completing 3 doses within 365 days or remaining partially vaccinated. Partial-vaccination clones were censored upon a third dose, and completion clones were censored at day 365 if a third dose had not been received. Follow-up ended at the first outcome, death, 5 years, study end, or strategy-specific artificial censoring. The composite outcome was persistent high-risk HPV infection (2 positive tests at least 6 months apart), cervical intraepithelial neoplasia grade 2 or higher, or cervical cancer [6].

We estimated clone-specific stabilized IPCW using baseline and time-varying covariates [7]. These weights addressed artificial censoring. Administrative censoring was assumed noninformative and was not separately weighted.

We fitted weighted Cox models with assigned strategy as the predictor and interpreted the hazard ratio (HR) as a model-based per-protocol contrast. Under conditional exchangeability of artificial censoring and correct IPCW models, no additional outcome-model covariates were included. The primary variance estimator was a robust sandwich estimator clustered by individual [8]. We compared this result with a naive Cox model, unclustered variance, and bootstrap at the individual level before cloning [9]. Sensitivity analyses used unstabilized weights and random forest (RF) censoring models.

## 3 Results

Among 3902 vaccine initiators, 923 (23.7%) completed 3 doses within 12 months and 2979 (76.3%) remained partially vaccinated, with 14 and 24 outcome events, respectively. Individuals who completed the series were older (mean [SD] age: 21.0 [3.3] vs 19.3 [3.3] years). The partially vaccinated group included larger proportions of non-Hispanic Black individuals (23.7% vs 11.2%) and individuals who initiated vaccination during 2021-2023 (30.5% vs 23.5%).

In the naive analysis, vaccine completion was associated with a higher hazard of HPV-related cervical disease (HR = 1.85, 95% CI: 0.95-3.57; Table 1). After CCW with unclustered variance, the association reversed (HR = 0.44, 95% CI: 0.18-1.04). Naive and CCW-weighted Kaplan– Meier curves are shown in Multimedia Appendix Figure S1. Individual-clustered sandwich variance left the HR unchanged but yielded a narrower interval (HR = 0.44, 95% CI: 0.33-0.57). The individual-level bootstrap produced a similar interval (HR = 0.44, 95% CI: 0.34-0.62) but required approximately 45 minutes for 1,000 iterations.

**Table 1:**
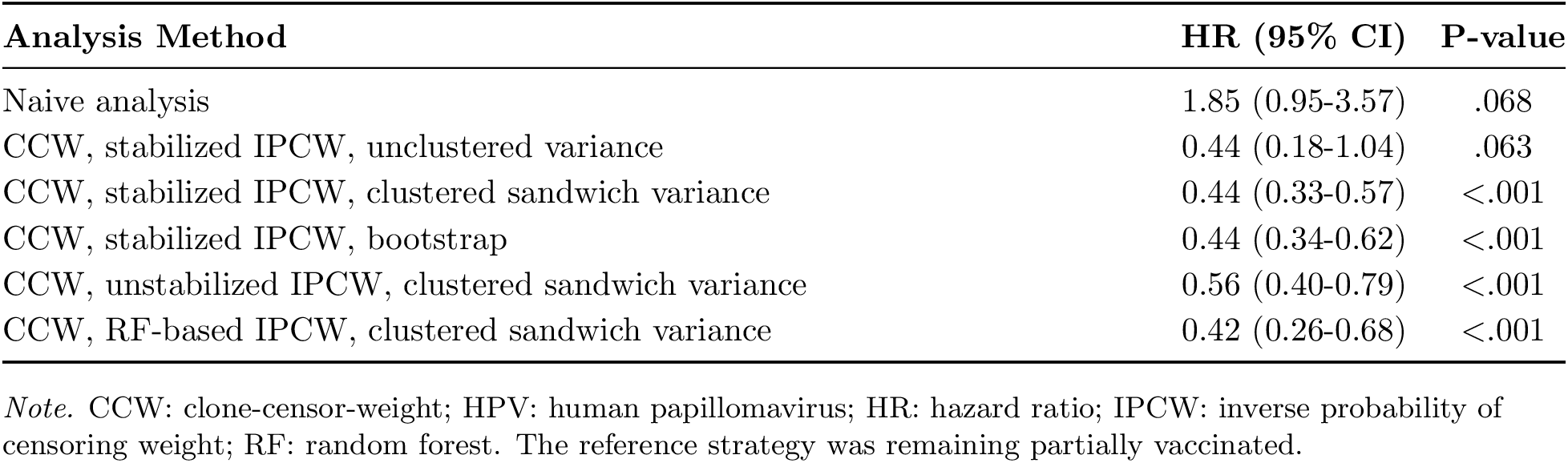
Association of completing the 3-dose 9vHPV series within 12 months with HPV-related cervical disease.

| Analysis Method | HR (95% CI) | P-value |
| --- | --- | --- |
| Naive analysis | 1.85 (0.95-3.57) | .068 |
| CCW, stabilized IPCW, unclustered variance | 0.44 (0.18-1.04) | .063 |
| CCW, stabilized IPCW, clustered sandwich variance | 0.44 (0.33-0.57) | <.001 |
| CCW, stabilized IPCW, bootstrap | 0.44 (0.34-0.62) | <.001 |
| CCW, unstabilized IPCW, clustered sandwich variance | 0.56 (0.40-0.79) | <.001 |
| CCW, RF-based IPCW, clustered sandwich variance | 0.42 (0.26-0.68) | <.001 |

*Note.* CCW: clone-censor-weight; HPV: human papillomavirus; HR: hazard ratio; IPCW: inverse probability of censoring weight; RF: random forest. The reference strategy was remaining partially vaccinated.

Stabilized weights were mostly concentrated below 15 without extreme values. The sensitivity analysis with unstabilized weights yielded a higher estimate (HR = 0.56, 95% CI: 0.40-0.79). Estimates from random forest weight models varied with tuning. Minimum leaf sizes of 20, 50, and 100 yielded HRs of 0.11, 0.18, and 0.30, respectively, and a leaf size of 200 yielded the estimate closest to the primary result (HR = 0.42, 95% CI: 0.26-0.68).

## 4 Discussion

The naive and CCW estimates pointed in opposite directions, illustrating how immortal time bias can reverse an estimated association when treatment is defined during follow-up. This work shows why CCW is a design-stage solution rather than a post hoc adjustment. Cloning represents each individual under every strategy at time zero, artificial censoring removes follow-up when observed treatment becomes incompatible with the assigned strategy, and IPCW reconstructs the strategy-specific risk sets under sustained adherence. Such framework distinguishes CCW from naive comparisons that use postbaseline treatment information to define groups.

Cloning also creates correlated records. Ignoring within-individual dependence can invalidate interval estimates even when the point estimate is unchanged. Here, individual-clustered sandwich variance and individual-level bootstrap methods produced similar intervals, whereas the unclustered interval led to a different inferential conclusion. Because bootstrapping requires resampling individuals before cloning and repeating the entire analysis, it is considerably more computationally intensive.

The model specification for weights remains important. Stabilized IPCW showed no extreme values, but estimates changed with unstabilized weights and across random forest specifications. Flexible models do not replace careful TTE design, weight diagnostics, or sensitivity analyses.

This EHR example shows how CCW prevents early risk time from being allocated according to future treatment information and handles strategy incompatibility transparently. Limitations include potential residual confounding and limited generalizability beyond a single health system. The weighted Cox HR is a model-based per-protocol contrast and should not, by itself, be interpreted as a causal effect. By connecting potential outcome reasoning with a reproducible analytic workflow, this study offers practical guidance for TTE with time-to-event outcomes and treatment strategies defined during follow-up.

## Data Availability

Access to the minimum dataset is available to qualified investigators through the UF Health IDR, subject to institutional review and data governance approvals.

